# The SARS-CoV-2 infection among students in the University of Porto: a cross-sectional study

**DOI:** 10.1101/2021.10.14.21264978

**Authors:** Paula Meireles, Joana Pinto Costa, Maria João Novais, Daniela Miranda, Mariana Mendes Lopes, Milton Severo, Henrique Barros

**Author notes:** equally contributing first authors (Paula Meireles, Joana Pinto Costa).

## Abstract

**Introduction:** Incidence based on notified cases of SARS-CoV-2 infection underestimates the real extension of the infection. We aimed to quantify SARS-CoV-2 specific antibodies’ seroprevalence among University students in Porto.

**Methods:** A rapid point of care testing for SARS-CoV-2 specific immunoglobulin (Ig) M and IgG antibodies was performed, and a questionnaire was applied to the 6512 voluntary students from September to December 2020. We computed the apparent IgM, IgG and IgM or IgG prevalence, and the true prevalence and 95% credible intervals (95% CI) using Bayesian inference.

**Results:** We found an apparent prevalence (IgM or IgG) of 9.7%, the true prevalence being 7.9% (95% CI 4.9-11.1). Prevalence was significantly higher among males (10.9% vs 9.2%), international students (18.1% vs 10.4% local vs 8.8% nationally displaced) and increased with age. Those with a known risk contact, that experienced quarantine, had symptoms, or a previous negative molecular test had a higher seroprevalence. Of the 91 (1.4%) students who reported a molecular diagnosis, 86.7% were reactive for IgM or IgG.

**Conclusion:** Based on immunological evidence infection was 5.6 times more frequent than if based on a molecular diagnosis. The higher seroprevalence among male, older, and international students emphasizes the importance of identifying particular groups.

## INTRODUCTION

The infection with the severe acute respiratory syndrome coronavirus 2 (SARS-CoV-2) can follow many distinct courses, with ominous outcomes mostly in the elderly population and no or few unspecific symptoms mainly among young and healthy individuals [1-3]. Real-time Polymerase Chain Reaction (RT-PCR) is the diagnostic “reference standard”, but testing strategies changed over the course of the epidemic and varied according to local logistic capacity. Thus, confirmed cases are a suboptimal indicator of the extent of SARS-CoV-2 infection, and the magnitude of undiagnosed infections can vary widely [4]. SARS-CoV-2 seroprevalence studies are critical to monitor the epidemic evolution in a population and to inform public health measures, such as vaccine allocation [5]. Those studies estimate the number of past infections higher than the number of RT-PCR confirmed cases [6, 7]. In the case of an emergent agent, it is assumed that all population is initially susceptible; therefore, the presence of specific antibodies provides good estimates of the cumulative incidence particularly if the infection provides long-term serological immunity.

In Portugal, the first case of coronavirus disease 2019 (COVID-19) was diagnosed on March 2, 2020 and on March 16 a nationwide schools closure was decreed affecting all education levels [8] – around 2 million students, more than 346 thousand from higher education [9]. Schools and universities fully reopened in mid-September 2020, providing an excellent opportunity to obtain data on the serum status of a large sample of university young adults exposed to highly varied risk contexts.

This study aimed to estimate SARS-CoV-2 specific antibodies’ seroprevalence and its determinants among students at the University of Porto (U.Porto), assessed between September and December of 2020.

## METHODS AND PARTICIPANTS

All undergraduate and postgraduate students from the U.Porto were sent an email by the University communication office to invite them to perform a rapid serological test for SARS-CoV-2 specific immunoglobulin (Ig) M and IgG antibodies. Along with this email, an information leaflet was sent. Participation was voluntary, and students scheduled their appointment according to their convenience. They were invited to answer a face-to-face questionnaire conducted by the trained researcher who performed the test, while waiting for the result.

The questionnaire included the following demographic and social questions: sex, age, living in usual residence (yes; no, usual residence in the country; no, usual residence abroad), faculty, history of contacts with a confirmed SARS-CoV-2 case since January 2020, history of being quarantined since January 2020, symptoms (then categorized as asymptomatic; paucisymptomatic: defined as having or having had one or two of the following symptoms: cough, dyspnea, odynophagia, headache, vomiting or nausea, diarrhea, fever, arthralgias, myalgia, asthenia; and symptomatic defined as having or having had at least three symptoms listed before, or dysgeusia or anosmia), ever being tested for SARS-CoV-2 infection, previous SARS-CoV-2 infection diagnosis, dates of diagnosis and recovery, self-perception of the probability of having been infected.

Data reported in this study refer to the period between September 24 to December 15, 2020, during which 6512 students (approximately 20% of the 32,443 students of U.Porto) self-selected to have a point of care serological test. The participants’ characteristics are presented in Table 1.

**Table 1:**
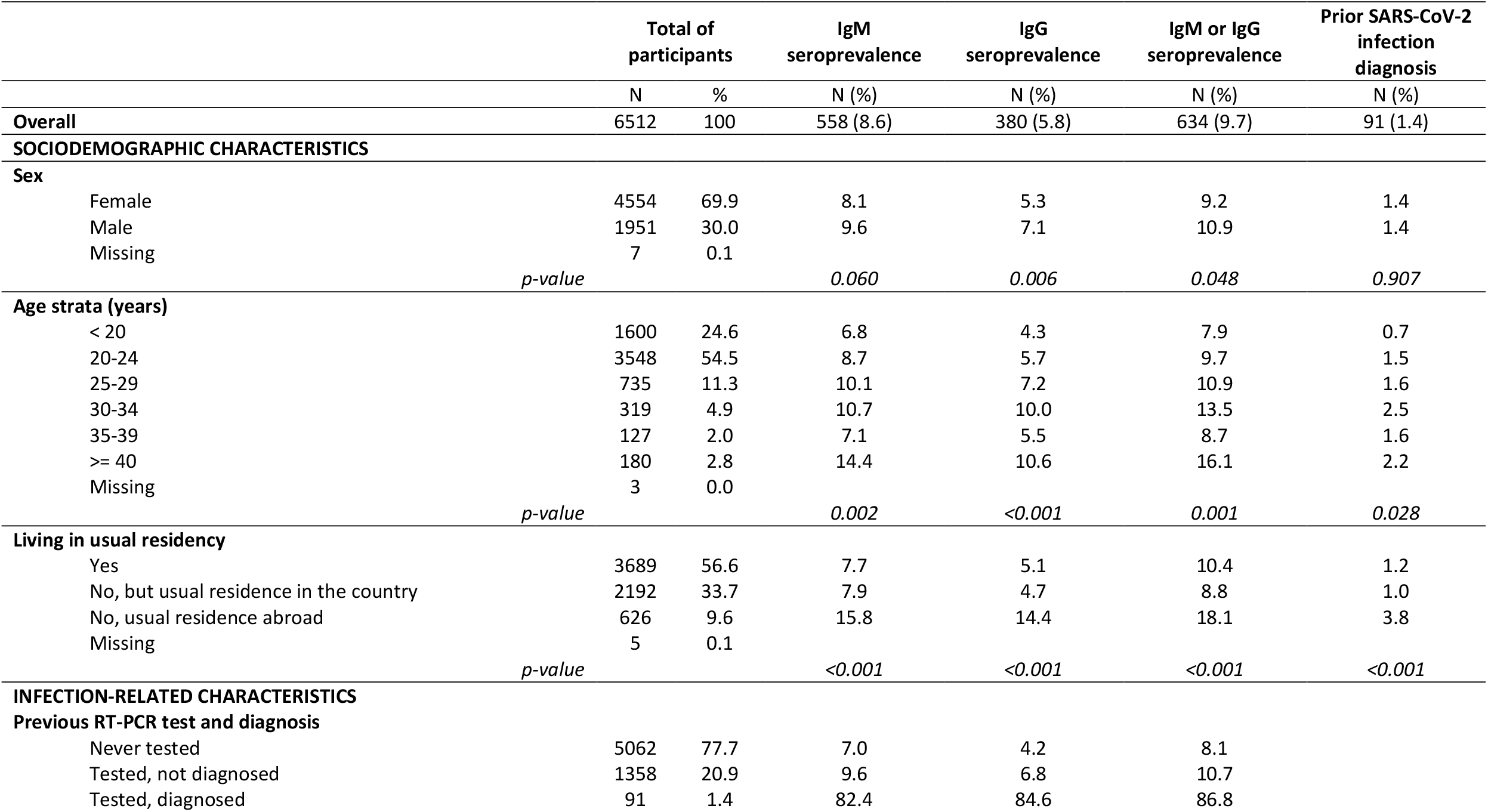
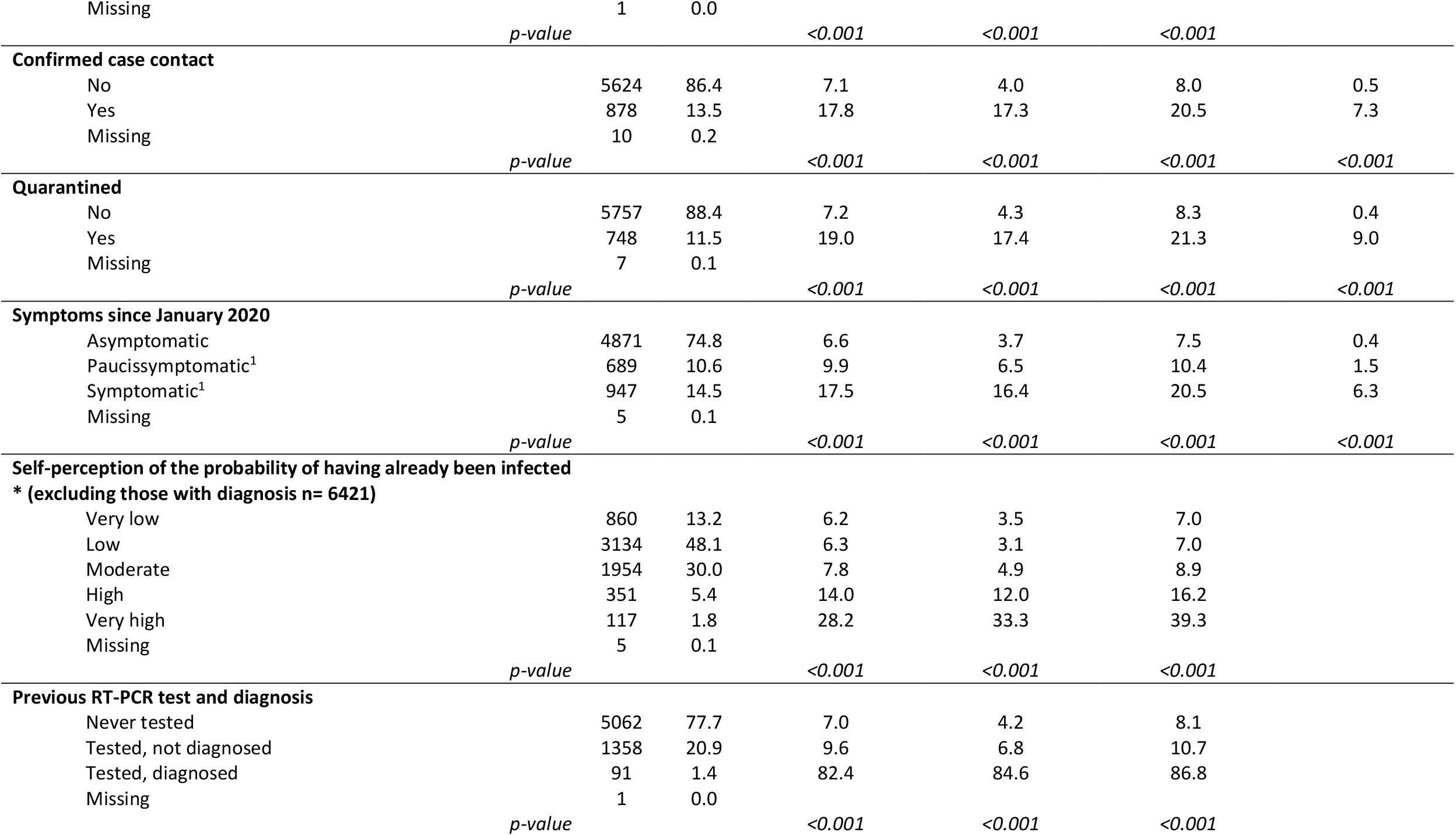
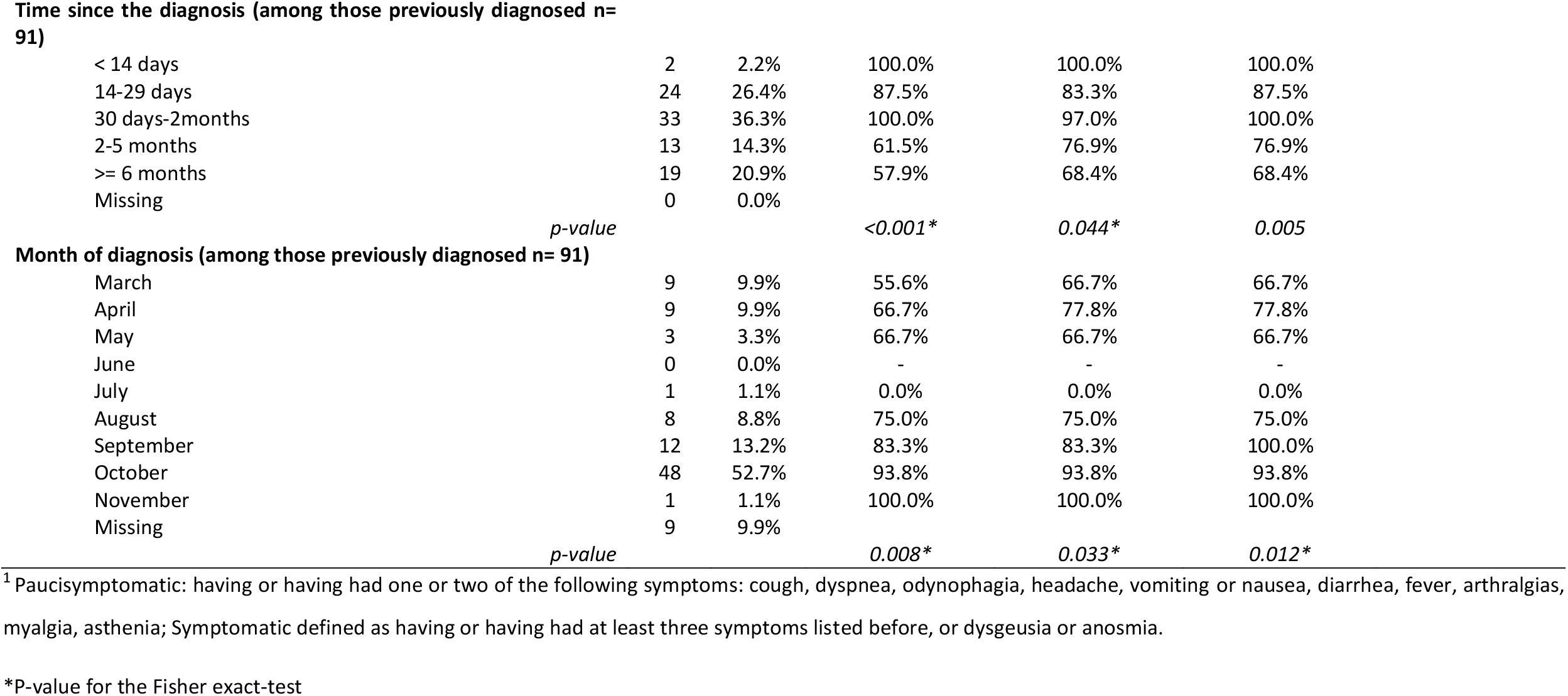
Characteristics of participants, IgM, IgG and IgM or IgG apparent seroprevalence and reported SARS-CoV-2 infection diagnosis by a molecular test according to those characteristics among U.Porto students from September to December 2020, Porto, Portugal

|  |  | Total of participants |  | IgM seroprevalence | IgG seroprevalence | IgM or IgG seroprevalence | Prior SARS-CoV-2 infection diagnosis |
| --- | --- | --- | --- | --- | --- | --- | --- |
|  |  | N | % | N (%) | N (%) | N (%) | N (%) |
| **Overall** |  | 6512 | 100 | 558 (8.6) | 380 (5.8) | 634 (9.7) | 91 (1.4) |
| **SOCIODEMOGRAPHIC CHARACTERISTICS** |  |  |  |  |  |  |  |
| **Sex** |  |  |  |  |  |  |  |
|  | Female | 4554 | 69.9 | 8.1 | 5.3 | 9.2 | 1.4 |
|  | Male | 1951 | 30.0 | 9.6 | 7.1 | 10.9 | 1.4 |
|  | Missing | 7 | 0.1 |  |  |  |  |
|  | *p-value* |  |  | 0.060 | 0.006 | 0.048 | 0.907 |
| **Age strata (years)** |  |  |  |  |  |  |  |
|  | < 20 | 1600 | 24.6 | 6.8 | 4.3 | 7.9 | 0.7 |
|  | 20-24 | 3548 | 54.5 | 8.7 | 5.7 | 9.7 | 1.5 |
|  | 25-29 | 735 | 11.3 | 10.1 | 7.2 | 10.9 | 1.6 |
|  | 30-34 | 319 | 4.9 | 10.7 | 10.0 | 13.5 | 2.5 |
|  | 35-39 | 127 | 2.0 | 7.1 | 5.5 | 8.7 | 1.6 |
|  | >= 40 | 180 | 2.8 | 14.4 | 10.6 | 16.1 | 2.2 |
|  | Missing | 3 | 0.0 |  |  |  |  |
|  | *p-value* |  |  | 0.002 | <0.001 | 0.001 | 0.028 |
| **Living in usual residency** |  |  |  |  |  |  |  |
|  | Yes | 3689 | 56.6 | 7.7 | 5.1 | 10.4 | 1.2 |
|  | No, but usual residence in the country | 2192 | 33.7 | 7.9 | 4.7 | 8.8 | 1.0 |
|  | No, usual residence abroad | 626 | 9.6 | 15.8 | 14.4 | 18.1 | 3.8 |
|  | Missing | 5 | 0.1 |  |  |  |  |
|  | *p-value* |  |  | <0.001 | <0.001 | <0.001 | <0.001 |
| **INFECTION-RELATED CHARACTERISTICS** |  |  |  |  |  |  |  |
| **Previous RT-PCR test and diagnosis** |  |  |  |  |  |  |  |
|  | Never tested | 5062 | 77.7 | 7.0 | 4.2 | 8.1 |  |
|  | Tested, not diagnosed | 1358 | 20.9 | 9.6 | 6.8 | 10.7 |  |
|  | Tested, diagnosed | 91 | 1.4 | 82.4 | 84.6 | 86.8 |  |
| Missing |  | 1 | 0.0 |  |  |  |  |
|  | *p-value* |  |  | <0.001 | <0.001 | <0.001 |  |
| **Confirmed case contact** |  |  |  |  |  |  |  |
| No |  | 5624 | 86.4 | 7.1 | 4.0 | 8.0 | 0.5 |
| Yes |  | 878 | 13.5 | 17.8 | 17.3 | 20.5 | 7.3 |
| Missing |  | 10 | 0.2 |  |  |  |  |
|  | *p-value* |  |  | <0.001 | <0.001 | <0.001 | <0.001 |
| **Quarantined** |  |  |  |  |  |  |  |
| No |  | 5757 | 88.4 | 7.2 | 4.3 | 8.3 | 0.4 |
| Yes |  | 748 | 11.5 | 19.0 | 17.4 | 21.3 | 9.0 |
| Missing |  | 7 | 0.1 |  |  |  |  |
|  | *p-value* |  |  | <0.001 | <0.001 | <0.001 | <0.001 |
| **Symptoms since January 2020** |  |  |  |  |  |  |  |
| Asymptomatic |  | 4871 | 74.8 | 6.6 | 3.7 | 7.5 | 0.4 |
| Paucissymptomatic<sup>1</sup> |  | 689 | 10.6 | 9.9 | 6.5 | 10.4 | 1.5 |
| Symptomatic<sup>1</sup> |  | 947 | 14.5 | 17.5 | 16.4 | 20.5 | 6.3 |
| Missing |  | 5 | 0.1 |  |  |  |  |
|  | *p-value* |  |  | <0.001 | <0.001 | <0.001 | <0.001 |
| **Self-perception of the probability of having already been infected**<br>**\* (excluding those with diagnosis n= 6421)** |  |  |  |  |  |  |  |
| Very low |  | 860 | 13.2 | 6.2 | 3.5 | 7.0 |  |
| Low |  | 3134 | 48.1 | 6.3 | 3.1 | 7.0 |  |
| Moderate |  | 1954 | 30.0 | 7.8 | 4.9 | 8.9 |  |
| High |  | 351 | 5.4 | 14.0 | 12.0 | 16.2 |  |
| Very high |  | 117 | 1.8 | 28.2 | 33.3 | 39.3 |  |
| Missing |  | 5 | 0.1 |  |  |  |  |
|  | *p-value* |  |  | <0.001 | <0.001 | <0.001 |  |
| **Previous RT-PCR test and diagnosis** |  |  |  |  |  |  |  |
| Never tested |  | 5062 | 77.7 | 7.0 | 4.2 | 8.1 |  |
| Tested, not diagnosed |  | 1358 | 20.9 | 9.6 | 6.8 | 10.7 |  |
| Tested, diagnosed |  | 91 | 1.4 | 82.4 | 84.6 | 86.8 |  |
| Missing |  | 1 | 0.0 |  |  |  |  |
|  | *p-value* |  |  | <0.001 | <0.001 | <0.001 |  |

**Time since the diagnosis (among those previously diagnosed n= 91)**
|  |  |  |  |  |  |
| --- | --- | --- | --- | --- | --- |
| < 14 days | 2 | 2.2% | 100.0% | 100.0% | 100.0% |
| 14-29 days | 24 | 26.4% | 87.5% | 83.3% | 87.5% |
| 30 days-2months | 33 | 36.3% | 100.0% | 97.0% | 100.0% |
| 2-5 months | 13 | 14.3% | 61.5% | 76.9% | 76.9% |
| >= 6 months | 19 | 20.9% | 57.9% | 68.4% | 68.4% |
| Missing | 0 | 0.0% |  |  |  |

*p-value*
*<0.001\**
*0.044\**
*0.005*

**Month of diagnosis (among those previously diagnosed n= 91)**
|  |  |  |  |  |  |
| --- | --- | --- | --- | --- | --- |
| March | 9 | 9.9% | 55.6% | 66.7% | 66.7% |
| April | 9 | 9.9% | 66.7% | 77.8% | 77.8% |
| May | 3 | 3.3% | 66.7% | 66.7% | 66.7% |
| June | 0 | 0.0% | - | - | - |
| July | 1 | 1.1% | 0.0% | 0.0% | 0.0% |
| August | 8 | 8.8% | 75.0% | 75.0% | 75.0% |
| September | 12 | 13.2% | 83.3% | 83.3% | 100.0% |
| October | 48 | 52.7% | 93.8% | 93.8% | 93.8% |
| November | 1 | 1.1% | 100.0% | 100.0% | 100.0% |
| Missing | 9 | 9.9% |  |  |  |

*p-value*
*0.008\**
*0.033\**
*0.012\**
<sup>1</sup> Paucisymptomatic: having or having had one or two of the following symptoms: cough, dyspnea, odynophagia, headache, vomiting or nausea, diarrhea, fever, arthralgias, myalgia, asthenia; Symptomatic defined as having or having had at least three symptoms listed before, or dysgeusia or anosmia.
\*P-value for the Fisher exact-test

The study protocol was approved by the ethics committee of the Institute of Public Health of the University of Porto (ID 20154). Verbal informed consent was obtained prior to the interview. Questionnaires were anonymous, and the results were only communicated to the students. The identifying information needed to schedule testing was kept only at the U.Porto information systems’ department. The linkage between datasets is impossible.

### SARS-CoV-2 specific IgM and IgG antibodies determination

Three point-of-care tests were used according to the manufacturer instructions – the STANDARD Q COVID-19 IgM/IgG Combo (manufacturer reported sensitivity of 94.5% seven or more days after symptom onset and specificity of 95.7% for both IgG and IgM), the HIGHTOP - SARS-CoV-2 IgM/IgG Test Combo (manufacturer reported sensitivity of 82.0% and 93.0% and specificity of 96.0% and 97.5% for IgM and IgG, respectively), and the *Teste Rápido Pantest de Coronavirus 2019-nCoV IgG/IgM* (manufacturer reported sensitivity of 85.0% and 100% and specificity of 96.0% and 98.0% for IgM and IgG, respectively). The three manufacturers used RT-PCR as the gold standard. The first was used from September 24 to October 19 (n=2263), the second from October 19 to 26 (n=1059), and the third from October 27 onwards (n=3190). All participants presenting with symptoms or reporting high-risk contacts in the previous 14 days were recommended to contact the National Health Service Contact Center. All participants were communicated their results orally and also in the form of a written leaflet with the information that the serological test only indicates whether there is evidence of previous contact with the SARS-CoV-2 and that it cannot be used to diagnose or rollout SARS-CoV-2 infection. It also recommended that all SARS-CoV-2 preventive measures were to be adopted and to call the National Health Service Contact Center in case of symptoms.

### Statistical analysis

We estimated seroprevalence as the proportion of individuals who had a reactive result in the IgM or IgG band of the point-of-care test. We estimated the true prevalence and 95% credible intervals (95% CI) using Bayesian inference. We used a uniform prior distribution for sensitivity ranging from 0.82 to 1 and specificity between 0.94 and 1. Estimates were obtained using the ‘rjags’ package in R.

Groups were compared using the Pearson Chi-Square, or the Fisher-exact test when the chi-square test’s assumptions did not hold.

## RESULTS

Table 1 presents the IgM, IgG and IgM or IgG apparent seroprevalence and reported SARS-CoV-2 infection prior diagnosis by a molecular test according to the characteristics of the U.Porto students. Among the 6512 students evaluated, 558 (8.6%) had a reactive test for IgM, 380 (5.8%) for IgG and 634 (9.7%) for IgM or IgG. The estimated true prevalence was 6.6 (95% CI 3.6-9.6) for IgM, 3.5 (95% CI 0.5-6.5) for IgG and 7.9 (95% CI 4.9-11.1) for IgM or IgG.

The prevalence of IgG was higher among males (7.1% vs 5.3% among females, p=0.006). The prevalence of IgM or IgG antibodies was higher among the 30-34 years old and the 40 and more years old, 13.5% and 16.1%, respectively (vs 7.9 % in students under 20, 9.7% in 20-24 years, 10.9% in 25-29 years and 8.7% in 35-39 years, p=0.001). A history of prior diagnosis was also higher in those age groups, 2.5% among the 30-34 years old and 2.2% in those aged 40 years or over (vs 0.7% in students under 20, 1.5% in 20-24 years, 1.6% in 25-29 years and 1.6% in 35-39 years, p=0.028).

The prevalence of antibodies was higher among international students (18.1% for IgM or IgG vs 10.4% among those living in their usual family home and 8.8% among nationally displaced, p<0.001). The proportion of those who report a previous infection diagnosis was also higher among international students (3.8% vs 1.2% among those living in their usual family home and 1.0% among nationally displaced, p<0.001).

Students who had contact with confirmed cases showed a prevalence of IgM or IgG of 20.5%, higher than the prevalence of 8.0% among those without (p<0.001). Similar results were found among those who were quarantined (21.3% vs 8.3%, p<0.001). IgM or IgG prevalence was also higher whenever there was a history of symptoms since the beginning of 2020, being 7.5% among asymptomatic, 10.3% among paucisymptomatic and 20.1% among ever symptomatic students (p<0.001).

SARS-CoV-2 infection had been previously diagnosed by a molecular test in 91 (1.4%) students. They had a prevalence of IgM or IgG antibodies of 86.8%; this was 10.7% in those tested negative and 8.1% in those never tested (p<0.001).

Of the 91 (1.4%) students who had been previously diagnosed with SARS-CoV-2 infection, 48 (52.7%) had a diagnosis in October 2020. The distribution by month of diagnosis showed an increase in cases starting in August 2020. The prevalence of antibodies decreased with the increasing time since diagnosis, 76.9% among those diagnosed between two and five months and 68.4% among those diagnosed six or more months before the serological test.

Among students without an infection diagnosis, the prevalence of antibodies increased with the increased perception of having been infected; it varied from 39.3% among those who considered this probability to be very high to 7.0% among those who thought it was low or very low.

## DISCUSSION

The 6512 students had a 9.7% prevalence of IgM or IgG antibodies. However, only 1.4% reported a prior diagnosis of SARS-CoV-2 infection based on an RT-PCR result. The burden of infection in this group was 6.9 times higher than the reported cases considering the point estimate or 5.6 times higher if compared with the estimated true prevalence of 7.9%, as observed in previously published surveys [4, 6, 7, 10]. The lower true prevalence was expected. Even using high specificity and sensitivity tests there is a high number of false positives due to the relatively low frequency of infection in this population [11].

Students had a higher prevalence of infection than observed in the Portuguese serological survey (ISNCOVID-19), conducted between May and July 2020 (2.9%) [12]. Considering the participants in the age group 20-39 years both in the ISNCOVID-19 and the U.Porto students, the prevalence was 2.9% and 10.1%, respectively. However, the studies were conducted in different periods of the epidemic in Portugal. The cumulative incidence of notified SARS-CoV-2 infection at the end of the national survey was 0.4% while at the end of this study, it was 3.5% [13]. These, along with differences in the recruitment of participants and the population’s characteristics, explain the difference in results.

Male and female students reported the same proportion of molecular diagnosis (1.4%). However, we found a higher seroprevalence among males, as reported in American university students [14], the Portuguese population [12] but not in other population-based surveys [4, 6, 7] and a meta-analysis [15]. We have no information on the study level (undergraduate or graduate) and therefore could not measure seroprevalence according to this variable, but older students had a higher seroprevalence. A previous study showed no difference in the prevalence of IgG antibodies in undergraduates and graduates suggesting they may have not had different lifestyles that would make them more or less susceptible [14] but considering age this may not have been the case in U.Porto. Students whose usual residence was abroad had a higher seroprevalence of infection. This might reflect a higher risk experience in their own countries or sharing a more vulnerable context during their stay in U.Porto.

We found higher seroprevalence among students who reported previous negative molecular test compared with those never tested, suggesting that some RT-PCR results might have been false negatives [16]. In accordance with previous studies, the self-reported belief of having had SARS-CoV-2 infection, prior contact with confirmed cases, and having had symptoms were positively associated with a higher seroprevalence [7, 12, 14].

Despite a previous RT-PCR positive test, 17.6% showed no IgM and 13.2% IgG antibodies. These may be false negative results, evidence of no immune response, or more likely waning of antibodies over time. The observed decreasing seroprevalence with increasing time after the diagnosis also support this explanation, as previously described [17, 18]. However, it is important to note that more than two-thirds of RT-PCR positive students had detectable immunological evidence of infection more than six months after diagnosis, indicating that antibodies may last long in a substantial proportion of individuals, as previously reported [19].

The national cumulative incidence of notified COVID-19 cases was 3.5% by the end of our data collection. However, only 1.4% of students reported a positive RT-PCR test for SARS-CoV-2.

This lower incidence may be partially explained by the higher proportion of young people with few or no symptoms and, therefore, unnoticed infections, students’ higher socioeconomic status, and an increased commitment to non-pharmacological preventive measures [1, 2]. It is worth mentioning that the number of reported cases among U.Porto students from September 16, 2020, to December 16, 2020, was 879, corresponding to 26 per 1000 students. This is almost double the observed in our sample and can have several explanations: 1. from an individual point of view those with previous infection may have less interest in doing the serological test because they know already they had contact with the virus; 2. they may be enrolled in care or having already an antibody test provided by the clinical services, and 3. some of those infections were recent and therefore students may have not yet had the opportunity to perform the test. We observed an increasing number of reported infection diagnosis in our study since August, as observed nationwide, but only one case in November, which might indicate that those diagnosed more recently did not yet have the chance to perform the test.

We used three different point-of-care tests over three different periods. This was unintended and was due to manufacturer delay on delivery which are constraints of real-world research in a time of high demand. However, all tests presented similar manufacturer’s reported characteristics, were used by the same trained researchers, and had similar performance in an in-house pilot test (data not shown). We cannot infer from this large sample to the U.Porto students’ population due to the lack of representativeness and the relatively low participation, probably explained by the voluntary nature of this testing program and the fact that many students remained on virtual learning. It is even possible that those who have had symptoms, high-risk contacts, or a self-perception of a higher probability of having been infected might be overrepresented. However, the dimension of infection in an educated, probably relatively low-risk community, is strong evidence of the increasing burden of COVID-19 in Portugal.

## Conclusion

In the University of Porto, students had a 7.9% seroprevalence, which was five times higher than the prevalence based on the reported molecular diagnoses and two times higher than the notified national cumulative incidence by the end of the study. Being an international student, reporting symptoms, self-perceiving high probability of infection, having had contact with a case, experiencing quarantine, and having had a diagnostic test performed though negative was associated with higher seroprevalence. Antibodies were present in 87% of those previously diagnosed with a molecular test, though reactivity decreased with increased time since the diagnosis.

## Data Availability

The datasets generated during and/or analyzed during the current study are available from the corresponding author on reasonable request.

## Funding

This study was funded by University of Porto and supported by national funds of Fundação para a Ciência e Tecnologia (FCT), under the scope of the project UIDB/04750/2020 - Research Unit of Epidemiology–Institute of Public Health of the University of Porto (EPIUnit). Joana Costa was the recipient of PhD grant 2020.08562.BD co-funded by the national funds of FCT and the Fundo Social Europeu (FSE). The funding source had no role in the work.

## Acknowledgements

We wish to acknowledge the team of researchers in the field namely Ana Margarida Lopes, Daniela Soares, Ema Fortunato, Flávia Mouta, Inês Roque, Inês Teixeira, Jacinta Mendonça, Janessa Oliveira, Laís Vieira, Mafalda Alves, Marta Costa, and Suellen Brito. The IT support from Paulo Oliveira, Celeste Pinto and Elisabete Neves. The U.Porto communication team led by Raul Santos. The U.Porto vice-rector Pedro Rodrigues.

## Competing interests

The authors have no conflicts of interest to declare that are relevant to the content of this article.

## Ethics approval

This study was performed in line with the principles of the Declaration of Helsinki. The study protocol was approved by the ethics committee of the Institute of Public Health of the University of Porto (ID 20154).

## Consent to participate

Verbal informed consent was obtained prior to the interview.

## Notes

### Competing Interest Statement

The authors have declared no competing interest.

### Funding Statement

This study was funded by University of Porto and supported by national funds of Fundacao para a Ciencia e Tecnologia (FCT), under the scope of the project UIDB/04750/2020 Research Unit of Epidemiology, Institute of Public Health of the University of Porto (EPIUnit). Joana Costa was the recipient of PhD grant 2020.08562.BD cofunded by the national funds of FCT and the Fundo Social Europeu (FSE). The funding source had no role in the work.

### Author Declarations

The Ethics Committee of the Institute of Public Health of the University of Porto gave ethical approval for this work (ID 20154).

